# An alarming high prevalence of resistance associated mutations to macrolides and fluoroquinolones in *Mycoplasma genitalium* in Belgium: Results from samples collected between 2015-2018

**DOI:** 10.1101/2020.03.22.20040840

**Authors:** I. De Baetselier, C. Kenyon, W. Vanden Berghe, H. Smet, K. Wouters, D. Van den Bossche, B. Vuylsteke, T. Crucitti

**Affiliations:** Institute of Tropical Medicine, Department of Clinical Sciences, STI Reference Laboratory, Antwerp, Belgium; Institute of Tropical Medicine, Department of Clinical Sciences, STI Unit, Antwerp, Belgium; Sciensano, Epidemiology of Infectious Diseases, Brussels, Belgium; Institute of Tropical Medicine, Department of Public Health, HIV/STI Unit, Antwerp, Belgium; Centre Pasteur du Cameroun, Yaoundé, Cameroun

**Keywords:** Mycoplasma genitalium, antimicrobial resistance, Belgium, fluoroquinolones, macrolides

## Abstract

**Objectives:** The number of reported cases of multi-resistant *Mycoplasma genitalium* (Mg) is increasing globally. The aim of this study was to estimate the prevalence of macrolide and possible fluoroquinolone resistance associated mutations (RAMs) of Mg in Belgium.

**Methods:** The study was performed retrospectively on two sets of Mg positive samples collected in Belgium between 2015-2018. The first set of samples originated from routine surveillance activities and the second set came from a cohort of men who have sex with men (MSM) using pre-exposure prophylaxis to prevent HIV transmission. Detection of RAMs to macrolides and fluoroquinolones was performed on all samples using DNA sequencing of the 23S ribosomal RNA gene, the *gyrA* gene and the *parC* gene.

**Results:** Seventy-six percent of the Mg samples contained a mutation conferring resistance to macrolides or fluoroquinolones (*parC* position 83/87). RAMs were more frequently found among men compared to women for fluoroquinolones (28.7% vs 9.1%) and macrolides (82.9% vs 27.3%). More than 90% of the Mg infections among MSM possessed a RAM to macrolides (90.7%). In addition, 23.4% of the samples harboured both macrolides and fluoroquinolone RAMs; 3.0% in women and 30.7% in MSM. Being MSM was associated with macrolide RAMs (Odds ratio: 15.1), fluoroquinolone RAMs (Odds ratio: 3.4) and having a possible multi-resistant Mg infection (Odds ratio: 8.2).

**Conclusion:** The study shows an alarmingly high prevalence of Mg with RAMs to macrolides and fluoroquinolones in Belgium. These results highlight the need to improve antimicrobial stewardship in Belgium in order to avoid the emergence of untreatable Mg.

**KEY MESSAGES:**

- Our study reports an alarmingly high prevalence of *Mycoplasma genitalium* antimicrobial resistance to either macrolides or fluoroquinolones in Belgium (76.6%).
- Multi-resistant Mg was found in almost 25% of cases, however, the prevalence of multi-resistant Mg was remarkably higher in men (27.1% vs 3.0%).
- The number of Mg resistant cases was much higher in men who have sex with men compared to heterosexual men and women.
- These results highlight the need to improve antimicrobial stewardship in Belgium.

## INTRODUCTION

*Mycoplasma genitalium* (Mg) is an emerging sexually transmitted infection (STI) and its global burden of disease is not very well known so far.[1]

Prevalence varies among different risk populations – and is generally higher in men who have sex with men (MSM) and female sex workers compared to other heterosexual individuals. A systematic review and meta-analysis up to 2017 found the prevalence of Mg to be 0.9% in pregnant women, 3.2% in MSM and 15.9% in female sex workers.[2] The prevalence of Mg in MSM, is, however, now reaching 10% according to the most recent publications.[3–6]

Mg is a cause of non-gonococcal urethritis in men and cervicitis in women, but Mg is also frequently found in asymptomatic individuals.[5] The natural history and clinical consequences of asymptomatic Mg infections are still poorly understood and including Mg in screening programmes among high risk groups is, therefore, not recommended. Furthermore, along with *Neisseria gonorrhoeae* (Ng), Mg is one of the STIs that can acquire resistance to different classes of antibiotics at an alarmingly rapid rate.[1] In fact, Mg cases with macrolide resistant associated mutations (RAMs) have been reported globally, and prevalence of these mutations is exceeding 80% in MSM using pre-exposure prophylaxis (PrEP) to prevent HIV.[3,4]

As a result, guidelines now suggest only testing and treating for Mg in cases of non-gonococcal, non-chlamydia urethritis or cervicitis. Moreover, Mg positive samples should be further analysed for the presence of macrolide resistance RAMs to tailor patient treatment and management.[7]

European guidelines recommend the use of moxifloxacin, a fourth generation fluoroquinolone, in cases where macrolide treatment failure is detected. [7] The prevalence of resistance to fluoroquinolones has been noted to be increasing in a number of regions. A recent systematic review from Europe estimated that the prevalence of fluoroquinolone resistance was 5% in this region, but there were considerable gaps in the data, such as from Belgium.[8]

With this study, we aimed to estimate the prevalence of RAMs to macrolides and fluoroquinolones in Mg samples received by the Belgian National Reference Centre (NRC) from 2015-2018. In addition, we explored risk factors associated with the presence of RAMs in order to inform clinical and testing practice.

## METHODS

### Samples

Since 2015, genital, anorectal, or urine samples from patients with clinical suspicion of Mg could be sent to the NRC, located at the Institute of Tropical Medicine (ITM), Antwerp for Mg identification (hereafter referred to as surveillance samples). Socio-demographic and clinical data, such as sexual orientation, HIV status, presence of symptoms, and presence of other STIs is requested for each case.

Samples from a PrEP demonstration study that was ongoing at the ITM from 2015 to 2018 were also included. The Be-PrEP-ared study was a single site, open-label PrEP demonstration study which included 200 HIV negative MSM.[9–11] A total of 179 HIV negative individuals were followed up for a period of 18 months and STI screening, including Mg detection, was performed quarterly at the three anatomical sites (i.e., urethra, anorectum and pharynx).[10]

For this retrospective study, both surveillance and study samples from 01 January 2015 to 31 December 2018 were included.

### Laboratory procedures

Mg detection was routinely performed at the NRC with an accredited in-house RT-PCR that targets the *pdhD*-gene until 30 September 2018 [12]. After this, Mg detection was performed using the S-DiaMGTV multiplex kit, according to manufacturer’s instructions (Diagenode diagnostics, Seraing, Belgium). DNA extraction of all samples was performed using the Abbott *m*2000sp (Abbott Molecular Des Plaines, Illinois, USA). Sample remnants and DNA extracts were stored below - 20°C.

Sanger sequencing of the 23S rRNA, *parC* and *gyrA* genes was performed as described previously to detect the presence of RAMs to macrolides and fluoroquinolones.[13,14]

Mutations found in region V of the 23SrRNA gene are reported according to the *Escherichia coli* numbering. Mutations found in the *ParC* (nucleotides 164–483) and *gyrA* gene (nucleotides 172–402) are provided according to *Mycoplasma genitalium* numbering. Samples with a silent mutation (without an alteration in amino acid) are not reported as mutations.

Mg was defined as resistant to macrolides if a RAM in the 23S rRNA region V was detected. Mg possessing an alteration in the *parC* gene at position 83 or 87 was defined as resistant to fluoroquinolones. Mg was defined as multi-resistant if mutations in both genes (23S rRNA and *parC* position 83/87) were detected.

### Statistical analysis

All data analyses were performed using STATA v.15.1.

The relationship between relevant socio-demographic and behavioural variables, and the presence of antimicrobial resistance to macrolides, fluoroquinolones, or both, was explored on all samples using two-tailed Fisher’s exact test.

If a relationship was detected, we report unadjusted and adjusted odds ratios (OR) with their corresponding 95% confidence intervals (CI). Variables significantly associated with the resistance of interest were selected for the multivariate logistic regression.

For all analyses, a significance level of 0.05 was used.

### Ethics statement

The Be-PrEP-ared study was approved by the Ethics Committee of the University of Antwerp and the Institutional Review Board of the ITM. All individuals provided informed consent.

Supplementary ethics approval was obtained for the detection of RAMs in Mg positive samples for both the Be-PrEP-ared study and the surveillance samples.

## RESULTS

### Description of the study population

Mg infections were found in 212 patients. The socio-demographic characteristics of the patients are described in Table 1. One third of the patients were included in the Be-PrEP-ared study (33.0%; n=70/212). Around one quarter of the patients had more than one positive Mg result over the four years (22.6%; n=48/212), most of them (79.1%; n=38/48) were part of the Be-PrEP-ared study.

**Table 1:** Socio-demographic characteristics of the study population. MSM: Men who have sex with men

|  | All participants |  | Women |  | Men |  |
| --- | --- | --- | --- | --- | --- | --- |
|  | No | % | No (n=64) | % | No | % |
| **Age (IQR)** | 32 | 26-40 | 26 | 23-32.5 | 35 | 28-42 |
| **Source** |  |  |  |  |  |  |
| Surveillance | 142 | 67.0 | 64 | 100 | 78 | 52.7 |
| Be-PrEP-ared | 70 | 33.0 | 0 | 0 | 70 | 47.3 |
| **Place of Residence** |  |  |  |  |  |  |
| Brussels | 47 | 22.2 | 2 | 3.1 | 45 | 30.4 |
| Flanders | 128 | 60.4 | 36 | 56.3 | 92 | 62.2 |
| Wallonia | 37 | 17.5 | 26 | 40.6 | 11 | 7.4 |
| **MSM** |  |  |  |  |  |  |
| No | 80 | 37.7 | 64 | 100 | 16 | 10.8 |
| Yes | 97 | 45.8 | 0 | 0 | 97 | 65.5 |
| Unknown | 35 | 16.5 | 0 | 0 | 35 | 23.7 |
| **HIV positive** |  |  |  |  |  |  |
| No | 117 | 55.2 | 15 | 23.4 | 102 | 68.9 |
| Yes | 15 | 7.1 | 1 | 1.6 | 14 | 9.5 |
| Unknown | 80 | 37.7 | 48 | 75 | 32 | 21.6 |
| **Positive for *Mycoplasma genitalium* at more than one visit** |  |  |  |  |  |  |
| No | 164 | 77.4 | 63 | 98.4 | 101 | 68.2 |
| Yes | 48 | 22.6 | 1 | 1.6 | 47 | 31.8 |
| **Number of visits** |  |  |  |  |  |  |
| 2 | 27 | 12.7 | 1 | 100 | 26 | 55.3 |
| 3 | 7 | 3.3 |  |  | 7 | 14.9 |
| 4 | 6 | 2.8 |  |  | 6 | 12.8 |
| 5 | 1 | 0.5 |  |  | 1 | 2.1 |
| 6 | 1 | 0.5 |  |  | 1 | 2.1 |
| 7 | 6 | 2.8 |  |  | 6 | 12.8 |

The 212 patients contributed to a total of 316 Mg positive samples. Around one fifth of the samples were from women (20.6%; n=65/316) and half of the Mg infections (50.9%; n=161/316) were detected in the framework of the Be-PrEP-ared study. Almost 45% of the infections were asymptomatic (44.9%; n= 142/316), around one quarter symptomatic (27.5%; n= 87/316) and the presence of symptoms was unknown for the other cases. (27.5%; n= 87/316). Urethritis was the most prevalent symptom in men (78.9%; n=56/71). Women mostly documented lower abdominal pain, or vaginal discharge (68.8%; n= 11/16). Cervicitis symptoms were reported in one quarter of the symptomatic cases in women (25.0%;n=4/16). STI co-infections were absent in half of the cases (54.4%; n=172/316) and in 86 cases the information was unknown (27.2%; n=86/316). A quarter of the Mg cases reported STI-associated symptoms (27.5%; n=87/316) and of those 20 were co-infected with another STI (23.0%; n=20/87).

### Presence of possible resistance associated mutations

Sequencing for both macrolide and fluoroquinolone RAMs was successful for 214 samples. Identified mutations and their resulting amino acid changes are shown in Table 2 for the 23S rRNA, *gyrA* and *parC* gene.

**Table 2:** Presence of *Mycoplasma genitalium* macrolide and possible fluoroquinolone resistance associated mutations (RAMs) in Belgium. CI: confidence interval; a: 10 samples were PCR negative and could not be sequenced for the *gyrA* gene; b: Samples presented an additional mutation in the *parC* gene: S83I in three of the samples and S83N in two samples; c: Three silent mutations were not counted as RAM and are categorized as wild type Mg; d: 21 Samples had an additional silent mutation (H78H (C234T) e: One sample had an additional mutation P62S (C184T); f: Samples presented an additional silent mutation (16 H78H (C234T) and 1 N99N (C297T))

| RAMs detected | Polymorphism<br>(mutation) | All Samples |  | Female Samples |  | Male Samples |  |
| --- | --- | --- | --- | --- | --- | --- | --- |
|  |  | No<br>(n=214) | % (95%CI) | No.<br>(n=33) | % (95% CI) | No.<br>(n=181) | % (95% CI) |
| Macrolide RAMS (*E. coli* numbering) |  |  |  |  |  |  |  |
| Wild Type |  | 55 | 25.7 (20.0-32.1) | 24 | 72.7 (54.5-86.7) | 31 | 17.1 (11.9-23.4) |
| Mutations detected |  | 159 | 74.3 (67.9-80.0) | 9 | 27.3 (13.3-45.5) | 150 | 82.9 (76.6-88.1) |
| 23SrRNA gene region V |  |  |  |  |  |  |  |
|  | A2058G | 65 | 30.4 | 4 | 12.1 | 61 | 33.7 |
|  | A2058T | 8 | 3.7 | 1 | 3.0 | 7 | 3.9 |
|  | A2059G | 86 | 40.2 | 4 | 12.1 | 82 | 45.3 |
| Possible Fluoroquinolone RAMS |  |  |  |  |  |  |  |
| Wild Type |  | 129 | 60.3 (53.5-66.7) | 20 | 60.6 (43.3-75.6) | 109 | 60.2 (52.9-67.1) |
| Mutations detected |  | 85 | 39.7 (33.3-46.5) | 13 | 39.4 (24.4-56.7) | 72 | 39.8 (32.9-47.1) |
| gyrA gene (*M. genitalium* numbering) |  |  |  |  |  |  |  |
|  | Wild Type | 207<sup>a</sup> | 96.7 (93.4-98.7) | 32 | 97.0 (84.2-99.9) | 175 | 92.3 (92.9-98.8) |
|  | Missense mutations | 7 | 3.3 (1.3-6.6) | 1 | 3.1 (0.77-15.8) | 6 | 3.3 (1.2-7.1) |
|  | A96T (G286A) | 1<sup>b</sup> | 0.5 | 0 | 0 | 1 | 0.6 |
|  | V85I (G253A) | 2<sup>b</sup> | 0.9 | 1 | 3.1 | 1 | 0.6 |
|  | M69I (G207A) | 1 | 0.5 | 0 | 0 | 1 | 0.6 |
|  | M95I (G285C) | 2<sup>b</sup> | 0.9 | 0 | 0 | 2 | 1.1 |
|  | A79V (C236T) | 1 | 0.5 | 0 | 0 | 1 | 0.6 |
| parC gene (*M. genitalium* numbering) |  |  |  |  |  |  |  |
|  | Wild Type<sup>c</sup> | 131<sup>c</sup> | 61.2 (54.3-67.8) | 20 | 60.6 (42.1-77.1) | 111 | 61.3 (53.8-68.4) |
|  | Missense mutations | 83 | 38.8 (32.2-45.7) | 13 | 39.4 (22.9-57.9) | 70 | 38.7 (31.5-46.2) |
|  | Mutations at 83 or 87 | 55 | 25.7 (20.0-32.1) | 3 | 9.1 (1.9-24.3) | 52 | 28.7 (22.3-35.9) |
|  | S83I (G248T) | 38<sup>d</sup> | 17.8 | 1 | 3.0 | 37 | 20.4 |
|  |  | No<br>(n=214) | % (95%CI) | No.<br>(n=33) | % (95% CI) | No.<br>(n=181) | % (95% CI) |
|  | S83N (G248A) | 4 | 1.9 | 1 | 3.0 | 3 | 1.66 |
|  | S83R (A247C) | 1<sup>e</sup> | 0.5 | 0 | 0 | 1 | 0.6 |
|  | D87N (G259A) | 4 | 1.9 | 1 | 3.0 | 3 | 1.7 |
|  | D87Y (G259T) | 8 | 3.8 | 0 | 0 | 8 | 4.4 |
| **Missense mutations at other locations in *parC*** |  |  |  |  |  |  |  |
|  | D82N (G244A) | 1<sup>e</sup> |  | 0 | 0 | 1 | 0.6 |
|  | P62S(C184T) | 26<sup>f</sup> |  | 9 | 27.3 | 17 | 9.4 |
|  | A118E (C353A) | 1 |  | 1 | 3.0 | 0 | 0 |
|  | **Silent mutations\*** | 3 |  | 1 | 3.0 | 2 |  |
|  | G75G (G225A) | 1 |  | 1 |  | 0 |  |
|  | H78H (C234T) | 1 |  | 0 |  | 1 |  |
|  | V112V (G336A) | 1 |  | 0 |  | 1 |  |

More than 80% of all Mg infections (n=177/214; 82.7%; 95%CI: 77.0-87.5) presented with a possible RAM to macrolides or fluoroquinolones. This applied to almost 90% of the samples collected from men (87.3%; 95%CI: 81.5-91.8). Women were more likely to have no RAMs (42.4%) compared to men (12.7%) (OR: 5.06; 95%CI: 2.03-12.3; p<0.0001).

#### Macrolide RAMs

Mutations conferring macrolide resistance at nucleotide position 2058 or 2059 of the 23S rRNA gene were detected in almost 75% of the samples (74.3%; n=159/214).

#### All possible fluoroquinolone RAMs

A total of 85 samples (39.7%) presented with possible RAMs in the gyrA gene or the *parC* gene (Table 2). Only seven samples showed a mutation in the *gyrA* gene, and five of these samples possessed an additional mutation in the *parC* gene (amino acid position 83). In total, almost 40% of the samples had a mutation in the *parC* gene (38.8%; n=83/214), and of those, 66.3% (n=55/83) had a mutation at location 83 or 87. Besides these two positions, mutation P62S (C184T) was found in 31.3% (n=26/83) of the Mg cases with *parC* mutations, and although this alteration was found in both genders, it was more present in women (27.3% vs 9.4%).

#### Presence of RAMs to both antibiotics

Almost one third of the samples (31.3%; n= 67/214; 95%CI: 25.2-38.0) harboured possible RAMs to both macrolides and fluoroquinolones. A2058G/S83I was the most frequent combination (31.3%; n=21/67), followed by A2059G/S83I (19.4%; n=13/67) and A2058G/P62S (16.4%; n=11/67).

### Associations between antimicrobial resistance and socio-demographic determinants

Due to the unknown relevance of the mutations found in the *gyrA* and *parC* gene, the exploration of risk factors for having fluoroquinolone resistance or multi-resistance was conducted on samples which harboured a mutation in the *parC* gene at position 83 and 87.

The number of macrolide resistant Mg cases was significantly higher in men (82.9%; 95%CI: 76,6-88,1) than in women (27.3%; 95%CI: 13,3-45,5; p<0.0001) (Table 3).

**Table 3:** Socio-demographics and presence of the different *Mycoplasma genitalium* genotypes. Associations were calculated using Fisher’s exact test. STI: Sexually Transmitted Infection; MSM: Men who have sex with men * All samples from return visits were from men and almost all were samples from MSM (70/72)

|  | Total |  | Wild type |  | p-value | Macrolide resistant<br>(23SrRNA) |  | p-value | Fluoroquinolone resistant<br>(*parC* position 83/87) |  | p-value | Multi-resistant<br>(23SrRNA + *parC*) |  |  |
| --- | --- | --- | --- | --- | --- | --- | --- | --- | --- | --- | --- | --- | --- | --- |
|  | No. | % | No. | % (95% CI) |  | No. | % (95% CI) |  | No. | % (95% CI) |  | No. | % (95% CI) | p-value |
| **All samples** | **214** |  | **50** | **23.4 (17.9-29.6)** |  | **159** | **74.3(67.9-80.0)** |  | **55** | **25.7 (20.0-32.1)** |  | **50** | **23.4 (17.9-29.6)** |  |
| **Year** |  |  |  |  |  |  |  |  |  |  |  |  |  |  |
| 2015-2016* | 61 | 28.5 | 6 | 9.8 (3.7-20.2) |  | 55 | 90.2 (79.8-96.3) |  | 16 | 26.2 (15.8-39.1) |  | 16 | 26.2 (15.8-39.1) |  |
| 2017 | 88 | 41.1 | 18 | 20.5 (12.6-30.4) |  | 67 | 76.2 (65.9-84.6) |  | 28 | 31.8 (22.3-42.6) |  | 25 | 28.4 (19.3-39.0) |  |
| 2018 | 65 | 30.4 | 26 | 40 (28.0-52.9) | <0.0001 | 37 | 56.9 (44.0-69.2) | <0.0001 | 11 | 16.9 (8.8-28.3) | 0.113 | 9 | 13.8 (6.5-24.7) | 0.083 |
| **Source of sampling** |  |  |  |  |  |  |  |  |  |  |  |  |  |  |
| Surveillance | 101 | 47.2 | 40 | 39.6 (60.0-49.8) |  | 57 | 56.4 (46.2-66.3) |  | 17 | 16.8 (10.1-25.6) |  | 13 | 12.9 (7.0-21.0) |  |
| Be-PrEP-ared | 113 | 52.8 | 10 | 8.8 (4.3-15.7) | <0.0001 | 102 | 90.3 (83.2-97.4) | <0.0001 | 38 | 33.6 (25.0-43.1) | 0.007 | 37 | 32.7 (24.2-42.2) | 0.001 |
| **Gender** |  |  |  |  |  |  |  |  |  |  |  |  |  |  |
| Women | 33 | 15.4 | 22 | 66.7 (48.2-82.0) |  | 9 | 27.3 (13.3-45.5) |  | 3 | 9.1 (19.2-24.3) |  | 1 | 3.0 (0.8-15.8) |  |
| Men | 181 | 84.6 | 28 | 15.5 (10.5-21.6) | <0.0001 | 150 | 82.9 (76.6-88.1) | <0.0001 | 52 | 28.7 (22.3-35.9) | 0.017 | 49 | 27.1 (20.7-34.2) | 0.001 |
| **Sexual Orientation** |  |  |  |  |  |  |  |  |  |  |  |  |  |  |
| Hetero | 48 | 22.4 | 30 | 62.5 (47.4-76.0) |  | 16 | 33.3 (20.4-48.4) |  | 4 | 8.3 (2.3-20.0) |  | 2 | 4.2 (0.5-14.3) |  |
| MSM | 140 | 65.4 | 11 | 7.9 (4.0-13.6) | <0.0001 | 127 | 90.7 (84.6-95.0) | <0.0001 | 45 | 32.1 (24.5-40.6) | 0.001 | 43 | 30.7 (23.2-39.1) | <0.0001 |
| Unknown | 26 | 12.2 | 9 | 34.6 (17.2-55.7) |  | 16 | 61.5 (40.6-79.8) |  | 6 | 23.1 (9.0-43.6) |  | 5 | 19.2 (6.6-39.4) |  |
| **HIV status** |  |  |  |  |  |  |  |  |  |  |  |  |  |  |
| Negative | 151 | 70.6 | 25 | 16.6 (11.0-23.5) |  | 125 | 82.8 (75.8-88.4) |  | 42 | 27.8 (20.8-35.7) |  | 41 | 27.2 (20.2-35.0) |  |
| Positive | 15 | 7.0 | 0 | 0 | 0.130 | 14 | 93.3 (68.1-99.8) | 0.469 | 4 | 26.7 (7.8-55.1) | 1.000 | 3 | 20.0 (4.3-48.1) | 0.761 |
| Unknown | 48 | 22.4 | 25 | 52.1 (37.2-66.7) |  | 20 | 41.7 (27.6-56.8) |  | 9 | 18.8 (8.9-32.6) |  | 6 | 12.5 (4.7-25.2) |  |
| **Type of Visit** |  |  |  |  |  |  |  |  |  |  |  |  |  |  |
| Initial Visit | 141 | 65.9 | 45 | 31.9 (24.3-40.3) |  | 93 | 66.0 (57.5-73.7) |  | 28 | 19.9 (13.6-27.4) |  | 25 | 17.7 (11.8-25.1) |  |
| Return visit* | 73 | 34.1 | 5 | 6.9 (2.3-15.3) | <0.0001 | 66 | 90.4 (81.2-96.1) | <0.0001 | 27 | 37.0 (26.0-49.1) | 0.008 | 25 | 34.3 (23.5-46.3) | 0.01 |
| **Site of Infection** |  |  |  |  |  |  |  |  |  |  |  |  |  |  |
| Genital | 148 | 69.2 | 40 | 27.0 (20.1-34.9) |  | 103 | 69.6 (61.5-76.8) |  | 36 | 24.3 (17.7-32.1) |  | 31 | 20.9 (14.7-28.4) |  |
|  | No. | % | No. | % (95% CI) |  | No. | % (95% CI) |  | No. | % (95% CI) |  | No. | % (95% CI) | p-value |
| Anorectal | 62 | 29.0 | 9 | 14.5 (6.9-25.8) | 0.053 | 53 | 85.5 (74.2-93.1) | **0.016** | 18 | 29.0 (18.2-41.9) | 0.494 | 18 | 29.0 (18.2-41.9) | 0.217 |
| Pharyngeal | 3 | 1.4 | 0 | 0 |  | 3 | 100(29.2-100.0) |  | 1 | 33.3 (0.84-90.6) |  | 1 | 33.3 (0.84-90.6) |  |
| Unknown | 1 | 0.5 | 1 | 100(25.0-100.0) |  | 0 | 0 |  | 0 | 0 |  | 0 | 0 |  |
| **STI associated Symptoms** |  |  |  |  |  |  |  |  |  |  |  |  |  |  |
| No | 99 | 46.3 | 13 | 13.1 (7.2-21.4) |  | 86 | 86.9 (78.6-92.8) |  | 31 | 31.3 (22.4-41.4) |  | 31 | 31.3 (22.4-41.4) |  |
| Yes | 62 | 29.0 | 16 | 25.8 (15.5-38.5) | 0.057 | 44 | 71.0 (58.1-81.8) | **0.023** | 15 | 24.2 (14.2-36.7) | 0.373 | 13 | 21.0 (11.7-33.2) | 0.203 |
| Unknown | 53 | 24.7 | 21 | 39.6 (26.5-54.0) |  | 29 | 54.7 (40.4-68.4) |  | 9 | 17.0 (8.1-29.8) |  | 6 | 11.3 (4.3-23.0) |  |
| **STI co-infections** |  |  |  |  |  |  |  |  |  |  |  |  |  |  |
| No STI co-infection | 128 | 59.8 | 19 | 14.8 (9.2-22.2) |  | 107 | 83.6 (76.0-89.5) |  | 35 | 27.3 (19.8-35.9) |  | 33 | 25.8 (18.5-34.3) |  |
| Any STI co-infection | 35 | 16.4 | 3 | 8.6 (1.8-23.1) | 0.415 | 30 | 85.7 (69.7-95.2) | 1.000 | 12 | 34.3 (19.1-52.2) | 0.410 | 10 | 28.6 (14.6-46.3) | 0.829 |
| Unknown | 51 | 23.8 | 28 | 54.9 (40.3-68.9) |  | 22 | 43.1 (29.3-57.8) |  | 8 | 15.7 (7.0-28.6) |  | 7 | 13.7 (5.70-26.3) |  |
| *Chlamydia* | 17 | 7.9 |  |  |  |  |  |  |  |  |  |  |  |  |
| *Gonorrhea* | 14 | 6.5 |  |  |  |  |  |  |  |  |  |  |  |  |
| *Trichomoniasis* | 1 | 0.5 |  |  |  |  |  |  |  |  |  |  |  |  |
| *Syphilis* | 3 | 1.4 |  |  |  |  |  |  |  |  |  |  |  |  |

This difference in gender was also seen in fluoroquinolone resistant Mg cases (28.7% vs 9.1%, p=0.017), however, to a lesser extent. The prevalence of multi-resistant Mg cases was remarkably higher in men (27.1%; 95%CI; 20.7-34.2) than in women (3.0%; 95%CI: 0.8-15.8; p= 0.001). Furthermore, being MSM increased the prevalence of all antibiotic resistant Mg genotypes. Almost one third (30.7%; 95%CI: 23.2-39.1, p<0.0001) of the Mg samples from MSM were multi-resistant, and 9 out of 10 infections among MSM had a mutation in the 23S rRNA gene (90.7%; 95%CI: 84.6-95.0, p<0.0001). Being of male gender and not MSM markedly decreased the presence of macrolide (56.1%; n=23/41) and fluoroquinolone resistance (32.1%; n=7/41). The number of wild type Mg samples increased to 41.5% (n=17/41), whereby the number of multi-resistant Mg cases dropped to 14.6% (n=6/41) (data not in table).

Other factors associated with antimicrobial genetic determinants were: having a previous positive test result, anorectal site of infection and being asymptomatic. The latter two were only significant for the presence of macrolide resistance.

On the multivariate analyses, being MSM was the only factor associated with a higher probability of having an Mg infection with macrolide resistance (AOR: 15.1 95%CI: 2.95-77.47; p=0.001), quinolone resistance (AOR: 3.40 95%CI: 1.28-12.49; p=0.017) or having a multi-resistant Mg infection (AOR: 8.23; 95%CI: 1.83-36.94; p=0.006) (Table 4).

**Table 4:** Crude odds-ratio(OR) and adjusted odds ratio’s (AOR) for the different *Mycoplasma genitalium* (Mg) genotypes. Adjustment was made for year, MSM, any sexually transmitted infection (STI) associated symptoms and returned visit for the presence of macrolide resistant-associated mutations. For the other Mg genotypes (Fluoroquinolone and multi-resistant Mg samples) adjustment was made for MSM and return visit CI: Confidence intervals

| Variable | Macrolide resistant<br>(23SrRNA) |  |  | Fluoroquinolone resistant<br>(*parC* position 83/87) |  |  | Multi-resistant<br>(23SrRNA + *parC* position 83/87) |  |  |
| --- | --- | --- | --- | --- | --- | --- | --- | --- | --- |
|  | OR<br>(95% CI) | AOR<br>(95% CI) | p-value | OR<br>(95% CI) | AOR<br>(95% CI) | p-value | OR<br>(95% CI) | AOR<br>(95% CI) | p-value |
| YEAR | 0.39<br>(0.25-0.61) | 0.81<br>(0.37-1.79) | 0.605 | - | - | - | - | - | - |
| Being in Be-PrEP-ared study | 7.16<br>(3.29-16.47) | - | - | 2.50<br>(1.25-5.13) | - | - | 3.30<br>(1.57-7.24) | - | - |
| Male Gender | 12.90<br>(5.11-34.22) | - | - | 4.03<br>(1.17-21.42) | - | - | 11.88<br>(1.86-493.29) | - | - |
| MSM | 19.54<br>(7.94-48.77) | 15.1<br>(2.95-77.47) | **0.001** | 5.21<br>(1.73-21.04) | 3.40<br>(1.28-12.49) | **0.017** | 10.20<br>(2.45-89.83) | 8.23<br>(1.83-36.94) | **0.006** |
| Return Visit | 4.87<br>(2.01-13.46) | 1.12<br>(0.32-3.97) | 0.86 | 2.37<br>(1.20-4.66) | 1.75<br>(0.86-3.55) | 0.122 | 2.42<br>(1.20-4.86) | 1.58<br>(0.77-3.24) | 0.214 |
| Anorectal Mg infection | 2.56<br>(1.12-6.37) | - | - | - | - | - | - | - | - |
| Any STI associated Symptoms | 0.37<br>(0.15-0.89) | 1.67<br>(0.50-5.56) | 0.406 | - | - | - | - | - | - |

A sensitivity analysis on samples from the first visit only, showed no statistically significant differences in the prevalence of resistance and, in addition, similar odds ratios were obtained for the variables of interest.

## DISCUSSION

A recent systematic review of macrolide and fluoroquinolone resistance in Mg found that global macrolide resistance increased to 46.3% by 2017, whilst fluoroquinolone resistance remained stable at around 6%.[15] In Europe, the fluoroquinolone resistance prevalence is now estimated to be 5%.[8] We found considerably higher rates of resistance in Belgium. In fact, less than 25% of the Mg strains analysed were wild type. More than 20% had RAMs to both macrolides and fluoroquinolones, and this was particularly prevalent in MSM – 30.7%. Previous reports from Belgium reported a lower prevalence of macrolide RAMs among female sex workers (6.9%) and MSM (44%).[6,16] It should be noted that in both studies another technique than the gold standard Sanger sequencing was used.

While Mg resistance has been described as occurring in MSM, the prevalence of RAMs in low-risk populations has not often been studied. The present study shows that RAMs to macrolides or fluoroquinolones are also prevalent in lower risk populations, such as women (33.3%) and heterosexual men (58.5%), but to a lesser extent than in MSM (92.1%). This may be explained by the relatively high consumption of both macrolides and fluoroquinolones in the general population in Belgium.[17]

In MSM, nine out of ten Mg infections were resistant to macrolides. An important driver of macrolide use in high STI-prevalence populations, such as PrEP cohorts, are chlamydia and gonorrhoea infections [18], where single dose azithromycin (alone or in combination) is a recommended therapy.[19,20] Individuals with chlamydia or gonorrhoea are frequently co-infected with Mg (as was the case for 20% in our study), which increases the probability of bystander antimicrobial selection pressure and which, in part, may explain the high prevalence of macrolide RAMs found in MSM.[4,21,22]

The present study also shows that asymptomatic Mg infections are prevalent, and that most of them (86.9%) presented with RAMs for macrolides or fluoroquinolones. Therefore, we add our findings to the evidence that screening for Mg and treatment of asymptomatic Mg cases should be reconsidered.

Treatment options for symptomatic infections with possible combined macrolide and fluoroquinolone resistant Mg are extremely limited. Pristinamycin is listed as third-line treatment in the IUSTI treatment guidelines, however, this treatment is difficult to obtain in Belgium. The current European IUSTI Mg treatment guidelines advocate azithromycin as first-line therapy.[7] The high prevalence of macrolide RAM in Belgium suggests that this approach is suboptimal. An alternative approach would be to follow the Melbourne protocol of treating Mg infections with doxycycline for 7 days whilst awaiting test results for macrolide resistance.[23] In the absence of macrolide resistance, azithromycin is administered, but if macrolide resistance is found then sitafloxacin/moxifloxacin is given.

While this protocol resulted in high cure rates and very low risk of genesis of macrolide resistance, our study results show that almost all MSM had a mutation conferring macrolide resistance, which is in line with other results.[3–5,24] Therefore, the detection of fluoroquinolone RAMs seems more relevant than the detection of macrolide resistance in this population.

In contrast to the commercial availability of several assays detecting macrolide RAMs in Mg, assays detecting fluoroquinolone RAMs are sparse.[25] There are different mutations in the quinolone resistance determining region of the *parC* and *gyrA* gene that have been associated with fluoroquinolone resistance. [25–28] In our study, only seven samples presented with a *gyrA* mutation and five of them coincided with a *parC* mutation at amino acid position 83, which seems to lead to high level resistance.[29] The two other alterations found (M69I and A79V) have not yet been correlated with clinical resistance. Due to this low number of mutations, the role of mutations in *gyrA* seems of less importance in Mg. The *parC* gene is more susceptible for mutations; in our study, most of the mutations were found at position 83 and 87, the ‘hot spots’ for fluoroquinolone resistance (n=55/83; 66.2%).[14] Besides these two positions, an alteration at position 62 (P62S) in the *parC* gene was detected in 32.9% of the samples with fluoroquinolone RAMs. This alteration has been previously described, however, clinical resistance has not yet been determined.[26,27,29]

Our study has several limitations; Firstly, the sampling methodologies may have introduced biases towards populations at higher risk for antimicrobial resistance, such as MSM with higher consumption of antimicrobials. Secondly, microbiological cure with moxifloxacin despite mutations at position 83/87 are documented, therefore the degree of fluoroquinolone resistance may be overestimated.[30] Thirdly, an unknown number of the isolates were from patients with treatment failure, which would likely exaggerate the prevalence of RAMs. Fourthly, clinical data was lacking for many of the surveillance cases, and finally, no test-of-cure within 14 days of treatment was performed. It is therefore unclear whether the infection was new or persistent.

In conclusion, we found high rates of macrolide and possible fluoroquinolone RAMs in Mg. These results provide further motivation to promote antimicrobial stewardship in the general population, but particularly amongst high STI prevalence populations such as PrEP cohorts. New treatment guidelines are urgently required that incorporate genotypic resistance testing for macrolides and fluoroquinolones. Finally, because of the high number of multi-resistant cases found in this study, new antimicrobials are urgently needed to treat patients with Mg resistance to both macrolides and fluoroquinolones.

## Data Availability

The data supporting the findings of this publication are retained at the Institute of Tropical Medicine (ITM), Antwerp and will not be made openly accessible due to ethical and privacy concerns. According to the ITM research data sharing policy, only fully anonymised data can be shared publicly. The surveillance data are de-identified (using a unique patient code) but not fully anonymised and it is not possible to fully anonymise them due to the longitudinal nature of the data. Data can however be made available after approval of a motivated and written request to the ITM at. The ITM data access committee will verify if the dataset is suitable for obtaining the study objective and assure that confidentiality and ethical requirements are in place.

## ACKNOWLEDGEMENTS

We would like to thank the laboratory staff of the STI reference laboratory and Wendy Thys for the additional check of all meta data.

